# Association of Germline BRCA Pathogenic Variants with Diminished Ovarian Reserve: A Meta-Analysis of Individual Patient-Level Data

**DOI:** 10.1101/2021.02.13.21251672

**Authors:** Volkan Turan, Matteo Lambertini, Dong-Yun Lee, Erica T Wang, Florian Clatot, Beth Y Karlan, Isabelle Demeestere, Heejung Bang, Kutluk Oktay

**Author notes:** Corresponding Author: Kutluk Oktay, MD, PhD, FACOG, Address: Yale School of Medicine, 333 Cedar Street, New Haven, CT 06510, Public correspondence.

## Abstract

**Purpose:** To determine whether germline *BRCA* pathogenic variants (g*BRCA*) are associated with decreased ovarian reserve.

**Materials and Methods:** An individual patient-data meta-analysis was performed using 5 datasets on 828 evaluable women who were tested for g*BRCA*. Of those, 250 carried g*BRCA* while 578 had tested negative and served as controls. Of the women with g*BRCA*, four centers studied those affected with breast cancer (n=161) and one studied unaffected individuals (n=89). The data were adjusted for the center, age, body mass index, smoking and oral contraceptive pill use before the final analysis. Anti-mullerian hormone (AMH) levels in affected women were drawn before pre-systemic therapy.

**Results:** Mean ages of women with vs. without g*BRCA1/2* (34.1± 4.9 vs. 34.3± 4.8 years; p=0.48), and with g*BRCA1* vs g*BRCA2* (33.7± 4.9 vs. 34.6± 4.8 years; p=0.16) were similar. After the adjustments, women with g*BRCA1/2* had significantly lower AMH levels compared to controls (23% lower; 95% CI: 4-38%, p=0.02). When the adjusted analysis was limited to affected women (157 with g*BRCA* vs. 524 without, after exclusions), the difference persisted (25% lower; CI: 9-38%, p=0.003). The serum AMH levels were lower in women with g*BRCA*1 (33% lower; CI: 12-49%, p=0.004) but not g*BRCA*2 compared to controls (7% lower; CI: 31% lower to 26% higher, p=0.64).

**Conclusions:** Young women with g*BRCA* pathogenic variants, particularly of those affected and with g*BRCA1*, have lower serum AMH levels compared to controls. They may need to be preferentially counselled about the possibility of shortened reproductive lifespan due to diminished ovarian reserve.

**Context:** *Key objective:* DNA repair deficiency is emerging as a joint mechanism for breast cancer and reproductive aging. Recent studies showed that ovarian reserve maybe lower in women with *BRCA* pathogenic variants (g*BRCA)* due to DNA repair deficiency. However, clinical studies using the most sensitive serum ovarian reserve marker Anti-Mullerian-Hormone (AMH) provided mixed results. Given the heterogeneity of the data from clinical studies, we performed an individual patient data (IPD) meta-analysis to determine if g*BRCA* are associated with lower ovarian reserve.

*Knowledge generated:* g*BRCA* are associated with diminished ovarian reserve, as determined by serum AMH and this association is restricted to g*BRCA1*. This finding is firmer for affected women as this IPD meta-analysis predominantly studied those with breast cancer.

*Relevance:* Women with g*BRCA* may have shortened reproductive life span due to diminished ovarian reserve and should be proactively counseled for fertility preservation especially if faced with chemotherapy or delaying childbearing.

## Introduction

*BRCA1* and *BRCA2* (*BRCA1/2*) play an essential role in double-strand DNA break (DSB) repair through recombination with undamaged, homologous DNA strands.^1^ Mutations in these genes are associated with increased susceptibility to breast and ovarian cancer.^2^ Starting with our first clinical and laboratory observations,^3-7^ a growing body of laboratory, translational and clinical evidence has emerged within the last decade, indicating a role for *BRCA* and related DNA DSB repair genes in ovarian function and aging.^6,7^

Anti-Mullerian hormone (AMH) is the best available serum marker of ovarian reserve. It is produced by granulosa cells of small antral and preantral follicles in the ovary, and by proportion, reflects the primordial follicle reserve.^8^ Serum AMH levels do not significantly fluctuate and can be measured at any point during the menstrual cycle. In contrast, the levels of indirect and less sensitive ovarian reserve markers such as FSH and E2 are highly dependent on the menstrual cycle day. One limitation for all ovarian reserve markers is that their levels can be affected by smoking, oral contraceptive use and obesity.^9^ Several studies have utilized serum AMH to investigate whether Germline *BRCA* Pathogenic Variants (g*BRCA)* are associated with diminished ovarian reserve. While a majority of studies indicated diminished ovarian reserve in women with g*BRCA1/2*, some provided conflicting results.^5-19^ Several clinical studies including our own,^5^ and transgenic mouse data indicated a stronger association of g*BRCA*1 with diminished ovarian reserve than with g*BRCA*2, however, one study found g*BRCA*2 but not the g*BRCA*1 to be associated with lower ovarian reserve.^15^ Several other studies did not detect lower serum AMH levels in women with g*BRCA* compared to controls.^14,18,19^

We recently performed a systematic review to investigate the role of g*BRCA* on ovarian aging.^6^ We found that the small sample size, lack of adjustment for important covariates (such as age, smoking and oral contraceptive pill use), not accounting for differences between the g*BRCA1* vs. g*BRCA2* carriers as well as inadequate statistical methods were among the major limitations of many studies investigating the association between g*BRCA* and serum AMH levels. To address these limitations and to provide more conclusive clinical assessment of this critical topic, we performed an individual patient-level data analysis with studies that investigated serum AMH levels in women with g*BRCA1/2*.

Based on laboratory^5^ and clinical data^6^, we hypothesized that AMH levels are lower in women with g*BRCA1/2*, especially in those carrying g*BRCA1*, compared to the individuals who tested negative for g*BRCA1/2*. To that end, we report the comparison of serum AMH levels in women with g*BRCA1/2* compared to those who were found to be negative for mutations in the same genes.

## Materials and methods

We searched for published articles in the PubMed database containing keywords, *BRCA, BRCA1, BRCA2*, mutations, *BRCA Pathogenic Variants*, ovarian reserve, Anti-Mullerian hormone in the English-language literature until December 2019. We found 12 original studies investigating the association between g*BRCA1/2* and serum AMH levels, four of which included only affected women with breast cancer ^5,10,20,21^, and one included both affected and unaffected ^18^ (Suppl. Table 1). After the study was approved by the Institutional Review Board (TR21092018/025), invitation letters were sent to all corresponding authors of the published articles. Four centers declined to participate and 3 did not respond. Of the seven non-participating centers, all studied unaffected women with the exception of one, which also included a small contingent of affected women. Five centers shared their individual patient-level data (IPD) from their publications. In addition, Lambertini et al. ^10^ updated their data with additional cases. In their published manuscript, the numbers of women with and without g*BRCA1/2* were 25 and 60 respectively. After updating their data, these numbers reached 50 and 85 respectively. As a result, a meta-analysis with five centers using IPD with some common key variables was conducted. Of those five centers, four (centers from NY/USA, South Korea, Belgium and France) studied women affected with breast cancer. One center studied unaffected women (Los Angeles/USA).

**Table 1.**
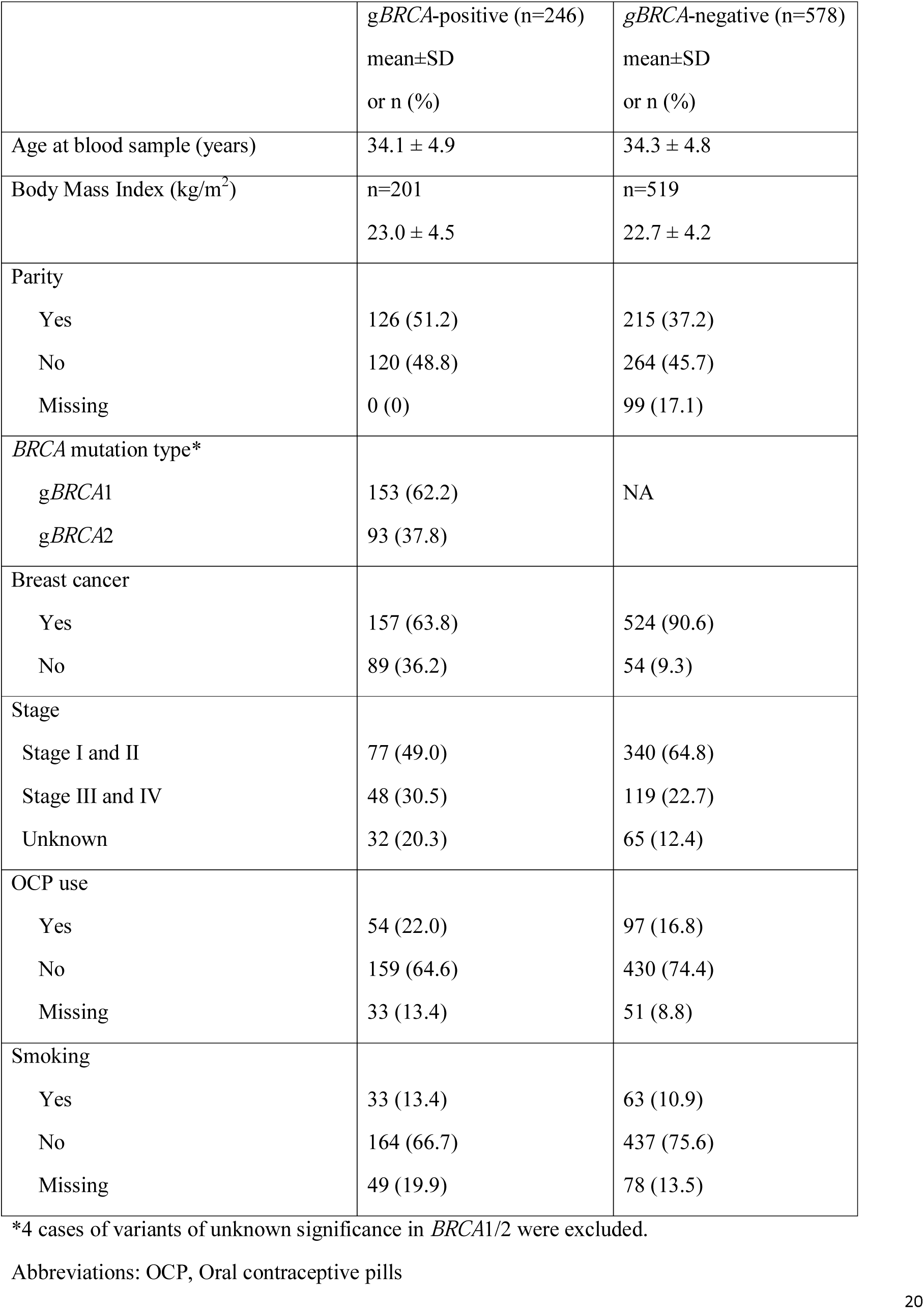
Population characteristics

|  | *gBRCA*-positive (n=246)<br>mean±SD<br>or n (%) | *gBRCA*-negative (n=578)<br>mean±SD<br>or n (%) |
| --- | --- | --- |
| Age at blood sample (years) | 34.1 ± 4.9 | 34.3 ± 4.8 |
| Body Mass Index (kg/m<sup>2</sup>) | n=201<br>23.0 ± 4.5 | n=519<br>22.7 ± 4.2 |
| Parity |  |  |
| Yes | 126 (51.2) | 215 (37.2) |
| No | 120 (48.8) | 264 (45.7) |
| Missing | 0 (0) | 99 (17.1) |
| *BRCA* mutation type* |  |  |
| *gBRCA1* | 153 (62.2) | NA |
| *gBRCA2* | 93 (37.8) |  |
| Breast cancer |  |  |
| Yes | 157 (63.8) | 524 (90.6) |
| No | 89 (36.2) | 54 (9.3) |
| Stage |  |  |
| Stage I and II | 77 (49.0) | 340 (64.8) |
| Stage III and IV | 48 (30.5) | 119 (22.7) |
| Unknown | 32 (20.3) | 65 (12.4) |
| OCP use |  |  |
| Yes | 54 (22.0) | 97 (16.8) |
| No | 159 (64.6) | 430 (74.4) |
| Missing | 33 (13.4) | 51 (8.8) |
| Smoking |  |  |
| Yes | 33 (13.4) | 63 (10.9) |
| No | 164 (66.7) | 437 (75.6) |
| Missing | 49 (19.9) | 78 (13.5) |

\*4 cases of variants of unknown significance in *BRCA1/2* were excluded.
Abbreviations: OCP, Oral contraceptive pills

**Table 2.** AMH values by overall g*BRCA* status (g*BRCA1* and g*BRCA2* combined). Pooled analysis with stepwise adjustment for center, age, smoking status, OCP use and BMI.

| Adjustment | % decrease in mean AMH (95% CI)<br>*gBRCA1/2* vs. none | P-value |
| --- | --- | --- |
| Center and age<br>(n=824) | 26 (9 to 39) | 0.004 |
| Center, age, smoking,<br>OCP<br>(n=824□) | 26 (9 to 39) | 0.004 |
| Center, age, smoking,<br>OCP, BMI<br>(n=720 □) | 23 (4 to 38) | 0.02 |
| Center, age, smoking,<br>OCP; only affected<br>women<br>(excluding Center-1)<br>(n=681) | 25 (9 to 38) | 0.003 |

□ We included missing data as a category for smoking and OCP, but did not impute missing BMI.
Abbreviations: AMH, Anti-Mullerian hormone; BMI, body mass index; OCP, Oral contraceptive pills

**Table 3.** AMH values by g*BRCA1* vs. g*BRCA2*. Pooled analysis with stepwise adjustment for center, age, smoking status, OCP use and BMI.

| Adjustment | % decrease in mean AMH (95% CI), adjusting age<br>*gBRCA1* vs. negative<br>*gBRCA2* vs. negative | P-value |
| --- | --- | --- |
| Center and age | 34 (17 to 49)<br>10 (32 to -19*) | 0.0004<br>0.47 |
| Center, age, smoking,<br>OCP | 35 (18 to 49)<br>9 (31 to -21) | 0.0004<br>0.52 |
| Center, age, smoking,<br>OCP, BMI | 33 (12 to 49)<br>7 (31 to -26) | 0.004<br>0.64 |
| Center, age, smoking,<br>OCP; only affected<br>women<br>(excluding center 1) | 32 (14 to 46)<br>14 (34 to -12) | 0.001<br>0.25 |

\*negative number means increase.
Abbreviations: AMH, Anti-Mullerian hormone; BMI, body mass index; OCP, Oral contraceptive pills

### Data collection and inclusion/exclusion criteria

For all participants enrolled in each of the included studies, IPD that contained demographics, parity, smoking status, oral contraceptive pill use, the g*BRCA1* and/or *gBRCA2* testing status, breast cancer stage (if affected) and serum AMH levels were collected. In affected women, serum AMH was drawn prior to the initiation of chemotherapy.

The common inclusion criteria were: Age 18–45, premenopausal status, no prior or ongoing chemotherapy or pelvic surgery, no use of endocrine therapy, and having been tested for g*BRCA1/2*. Other than one center (Los Angeles, USA), all excluded women with irregular periods and history of Polycystic Ovarian Diseases (PCOS) or other reproductive endocrine disorders.

### AMH assessment (please see Supplement 1)

#### Statistics

We summarized patient characteristics by g*BRCA* status using standard descriptive statistics – mean and SD for continuous variables and frequency and proportion for categorical variables. We set 0.01 as the detection limit and used log10-transformed AMH data following our previously published approach ^23^ and our examination of the AMH data for this IPD meta-analysis.

Data were analyzed using the statistical methods for multi-center studies or IPD meta-analysis with patient-level covariates and outcomes.^24-26^ The age-adjusted model was fit for 5 studies/centers individually, and sequentially-adjusted models (from ‘center and age only’ to ‘center, age, smoking, oral contraceptive pill use and body mass index) for the combined sample. Smoking and oral contraceptive pill (OCP) use were categorized to 3 levels (Y, N, missing) to avoid imputation and to utilize maximum sample size. Patients with missing body mass index (BMI) (as continuous variable) were not included when BMI was adjusted, i.e., we did not impute missing continuous variables, including BMI. Fixed effects models were chosen as the primary method as explained in Supplement 2.

The primary exposure of interest was g*BRCA* status (Y/N). In the secondary analysis, 3-level of g*BRCA* type 1 vs. g*BRCA* type 2 vs. negative (as reference group) were considered. We analyzed data using SAS 9.4 (SAS Institute, Cary, NC). AMH differences in each study and pooled version were visualized in a Forest plot.

## Results

### General description of the study population

After excluding 4 women with variance of unknown significance in *BRCA*, a total of 824 out of 828 women were eligible for the final analysis (Figure 1). Two hundred and forty-six women tested positive for g*BRCA*1/2 and 157 (78.5%) of those were affected with breast cancer. Eighty-nine women with g*BRCA* were unaffected. Of the 246 women with g*BRCA*, 153 (62.2%) were positive for g*BRCA*1 while 93 women (37.8%) for g*BRCA*2. Among the mutation negative controls (n=578), 524 were affected with breast cancer.

Women with and without g*BRCA* had similar age at study inclusion compared with non-carrier controls (mean±SD: 34.1±4.9 vs 34.3±4.8 years, respectively; p=0.48). The demographic characteristics of the entire/combined sample is summarized in Table 1.

**Figure 1.**
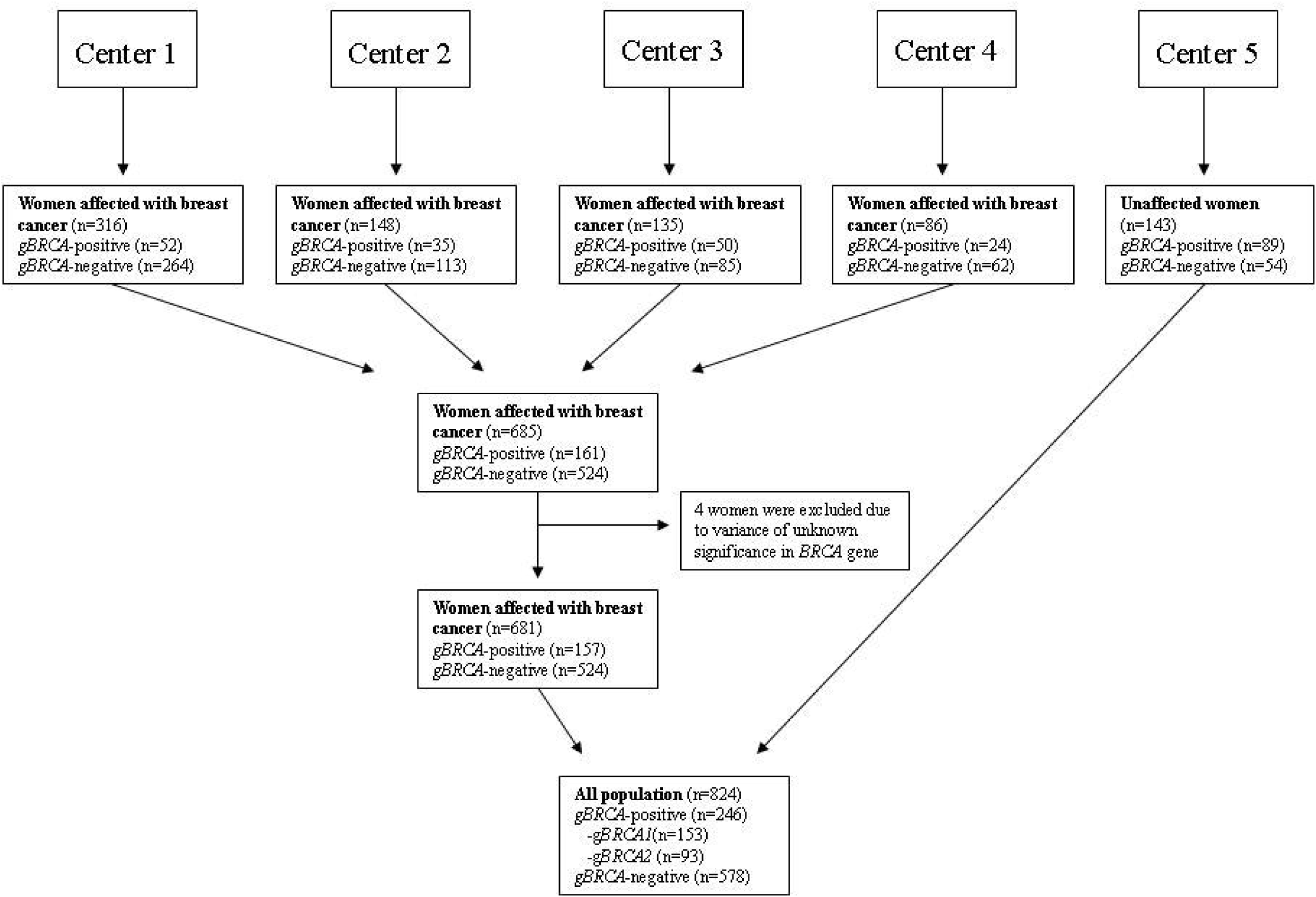
Study inclusion and exclusion flow chart.

### Comparison of serum AMH levels in women with and without gBRCA1/2

Mean AMH level in women with g*BRCA1/2* was 2.04 ng/ml (SD=2.0, median of 1.5, geometric mean of 0.99) while it was 3.36 ng/ml (SD=3.1, median of 2.5, geometric mean of 1.96) in women without mutations. After adjusting for center, age, smoking status and OCP use, women with g*BRCA* had significantly lower AMH levels compared with those without [26% lower (95% CI: 4-38%), p=0.004]. After the inclusion of BMI in the adjusted model, the sample size was reduced due to the missing data (824 to 720), but qualitatively similar results were observed; for example, we found 23% decrease in AMH (CI: 4-38%, p=0.02) for g*BRCA1/2* carriers vs. non-carriers (Figure 2).

**Figure 2.**
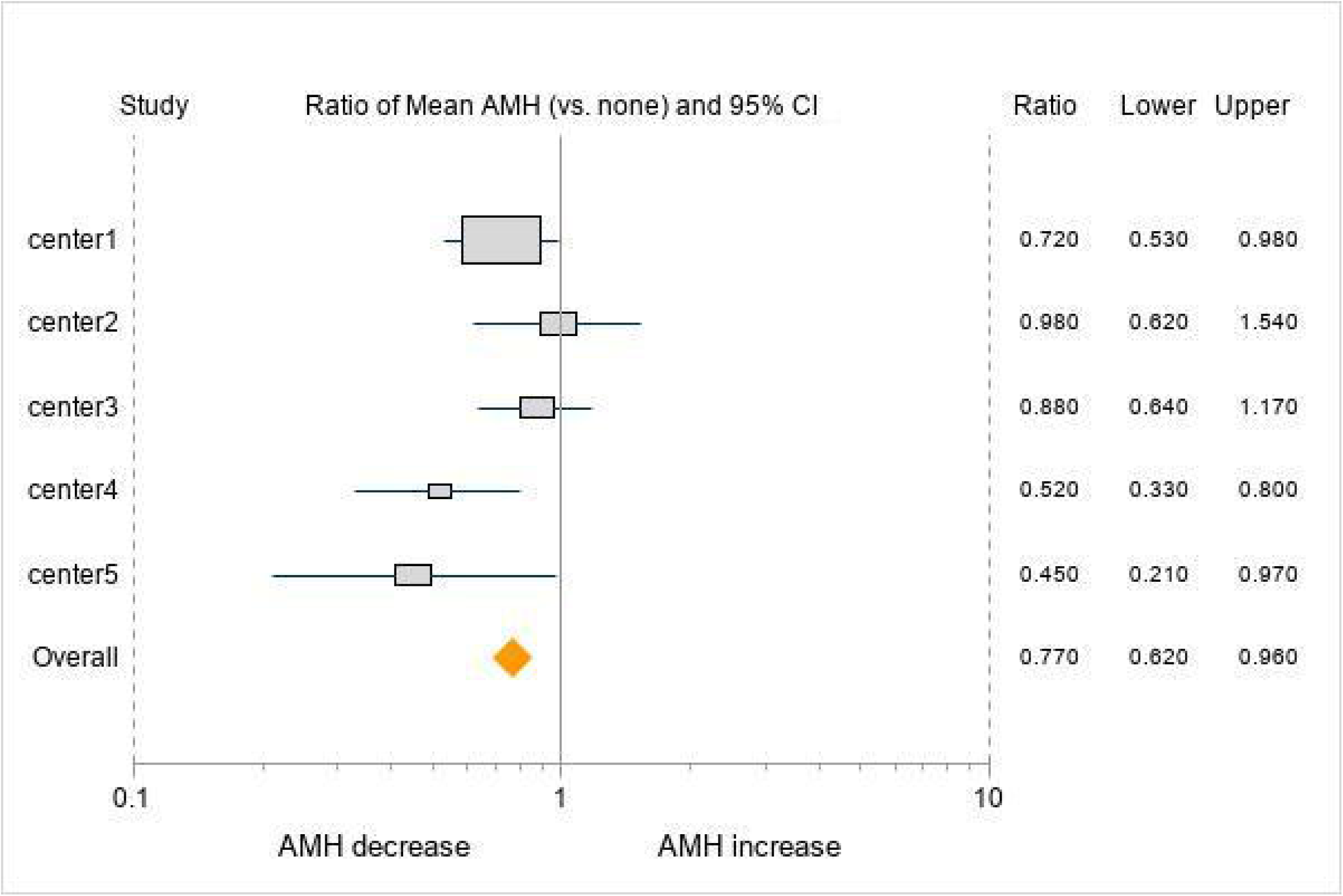
Forest plot analysis of individual results from five participating centers. 1 is null value of 0 difference or decrease. Individual centers were minimally adjusted, while overall data were adjusted for center, age, smoking, OCP and BMI.

### Comparison of serum AMH levels in women with gBRCA1 vs gBRCA2 to controls

To further evaluate whether there was a difference in ovarian reserve according to type of g*BRCA*, we categorized women into those with g*BRCA1*, g*BRCA2*, and no mutations. The comparison among these three groups showed that the AMH levels were significantly lower in women with g*BRCA1* compared to controls after adjusting for age, smoking and OCP use with (33% lower; p=0.004) or without (35% lower, p=0.0004) adjustment for BMI. A similar comparison of AMH levels between the women with g*BRCA2* and controls did not reveal a difference; 7% lower (p=0.64) and 9% lower (p=0.52), respectively.

### Comparison of serum AMH levels in affected (breast cancer) women with and without gBRCA

It is possible that the *BRCA* dysfunction is more severe in affected women with mutations and hence the ovarian reserve may be more severely compromised in the same group. Therefore, we repeated our analysis by excluding the data from Center-5 (n=143) which studied unaffected women with and without g*BRCA*. Of the remaining 681 women with a new diagnosis of breast cancer, 91 had g*BRCA1*, 66 had g*BRCA2*, and 524 tested negative for g*BRCA1/2*.

The mean ages of 157 and 524 women with and without g*BRCA*1/2 were 33.3±4.3 and 33.8±4.5 years respectively (p=0.18). Mean AMH level was 2.54 ng/ml (SD=2.3, median of 1.9, geometric mean of 1.66) in women with g*BRCA*, while it was 3.59 ng/ml in women without g*BRCA* (SD=3.2, median of 2.8, geometric mean of 2.31). After adjusting for center, age, smoking status, BMI and OCP use, women with g*BRCA* had significantly lower AMH levels compared with women without g*BRCA* [25% lower CI: 9-38%, p=0.003]. Furthermore, after adjusting for center, age, smoking and OCP use, AMH levels of women with g*BRCA1* were lower compared to the controls (32% lower, CI: 14-46% lower, p=0.001). The serum AMH levels of women with g*BRCA*2 showed no significant difference in comparison to controls (14% lower, CI: 34% lower −12% higher, p=0.25).

### Secondary/Sensitivity analysis with RE vs. FE

When we fitted an RE model as a secondary/sensitivity analysis, our analysis also showed robust results. For example, when we estimated % decrease in mean AMH between g*BRCA*1/2 vs. none, adjusting center and age only, our original analysis yielded 26 (95% CI: 9 to 39; p=0.004), while a newly fitted RE model yielded 27 (15 to 37; p≤0.0001). As another extreme case with only affected women, adjusting for center/age/smoking and OCP use, the estimated % decrease was 25 (9 to 38; p=0.003) vs. 25 (10 to 38; p=0.002) for these two models, respectively. This sensitivity analysis shows that, the FE model that we used for the primary analysis for our IPD data was slightly more conservative than the RE model.

## Discussion

We performed an individual patient-level data analysis from five centers to investigate the relationship between g*BRCA* and AMH levels. After adjusting for potential confounders, we found that women with g*BRCA*, specifically those affected and carrying g*BRCA1*, have lower serum AMH levels compared with women without g*BRCA*. To our knowledge, this is the first multicenter analysis and the largest study investigating the relationship between g*BRCA* and AMH levels in women with or at risk for breast cancer.

Oktay et al. first reported low response to ovarian stimulation ^3^, and subsequently, lower serum AMH levels in women with breast cancer.^5^ This was followed by several studies supporting the finding of lower serum AMH in both affected and unaffected women with g*BRCA* ^5,11,13,20^, but others were unable to detect similar differences.^14,18,19^ We have recently reviewed the published evidence on the impact of g*BRCA* on ovarian aging and discussed the limitations and possible reasons for discrepancies among the studies.^6^ This individual patient level meta-analysis of published and updated data was performed to overcome the shortcomings of distinct studies. The current study confirmed the findings from most studies that particularly the presence of a g*BRCA1* negatively impacts on the ovarian reserve of young women affected with or at risk for breast cancer.

Laboratory studies in human ovarian tissue have determined the potential mechanism of diminished oocyte reserve in women with g*BRCA. BRCA1/2* are the members of the ATM-mediated DNA double strand break (DSB) repair pathway. Inadequate repair of DNA DSBs results in severe mutagenesis leading to carcinogenesis and tissue aging.^27,28^ The ATM-mediated DNA DSB repair pathway is charged with repair of this most lethal form of DNA damage, which is estimated to occur nearly a million times every day.^27^ The basic research from Dr. Oktay’s laboratory showed that g*BRCA1* but not the g*BRCA2* mutant mice have fewer primordial follicles which accumulate more DNA DSBs in their oocytes with age compared to the wild type mice. These mice also ovulate fewer oocytes and have lower litter size than the controls.^5^

The same team has also shown that the ovaries of women with g*BRCA* carry fewer primordial follicles, which are lost at an accelerated fashion and accumulate more DNA DSBs with age compared to ovaries from controls.^12^ Oktay’s laboratory also showed that gonadotoxic chemotherapy induces primordial follicle death and ovarian reserve loss by inducing DNA DSBs and apoptosis of oocytes, and some oocytes may be able to repair themselves by activating the ATM-Pathway.^29^ In addition, recent longitudinal and laboratory data suggest that women with g*BRCA* may lose larger ovarian reserve after chemotherapy due to oocyte DNA repair deficiency.^23,30,31^ This may be a “double whammy” for affected women with gBRCA as their already lower ovarian reserve status is compounded by larger chemotherapy-induced loss, rendering them highly liable for infertility. However, further studies will be needed to confirm that women with gBRCA lose clinically significantly larger ovarian reserve after chemotherapy, compared to those without mutations.

In fact, *BRCA1* and other ATM-pathway genes and the age-induced decline in their function appear to be central in ovarian aging.^5,6^ *BRCA1* has a more complex involvement in the ATM-mediated DNA DSB repair pathway than *BRCA2*. While *BRCA1* plays a role in damage sensing, homologous recombination repair and check-point regulation (such as through CHEK2), the role of *BRCA2* is limited to homologous recombination only. Moreover, the age-related decline in the *BRCA1* function has been shown to occur earlier than the *BRCA2* function in human oocytes.^5^ While that decline appears to become prominent in the third decade of life in women with g*BRCA1*, the same may not happen until the fourth decade in the case of g*BRCA2*.^5^ Because women with g*BRCA* have one dysfunctional allele, age-induced decline in the function of the intact allele results in an acquired homozygocity with age.^31^ This then results in the accelerated accumulation of DNA DSBs in human oocytes, which trigger apoptotic death mechanisms of cell senescence, resulting in the accelerated reduction of ovarian reserve.^5,6,23^ Because the function of *BRCA1* declines earlier in life than the *BRCA2*, this may explain why ovarian reserve loss is more prominent in women with g*BRCA*1 compared to g*BRCA*2.

Considering the possibility that the affected women may have more severely accelerated ovarian aging, we analyzed our data with and without the inclusion of unaffected women, but this analysis did not alter our results. In our IPD analysis, only one center (Cedars Sinai Medical Center, LA, USA) studied unaffected women and found lower serum AMH in those with g*BRCA*1. In this meta-analysis, we included all published studies in affected women while the non-participating studies, except for one (Gunnala et al. studied both affected and unaffected women), were performed among the unaffected (Suppl. Table 1).^18^ In total, there have been six studies that assessed the relationship between g*BRCA1/2* and AMH levels only in unaffected women. While two studies were negative,^14,19^ four showed that there were lower AMH levels in women with g*BRCA1/2*^32^, with only g*BRCA1*^13,33^ or only g*BRCA2*.^15^ Therefore, while the preponderance of evidence also suggests lower serum AMH in unaffected women compared to controls, further research is needed to determine the magnitude of serum AMH differences between affected and unaffected women and those with *gBRCA1* vs. *gBRCA2*. Therefore, our findings are on firmer ground with affected women with g*BRCA*.

There is other evidence supporting lower ovarian reserve in women with g*BRCA*. Several studies showed earlier menopausal age, particularly for those with g*BRCA*1.^16,34^ A large meta-analysis of genome-wide associations analysis identified polymorphism in the *BRCA*1 as one of the key determinants of age at natural menopause.^35^

It is also possible that the differences we have reported here are underestimations as those most severely affected might have already had early risk reducing salpingo-oophorectomy and or developed early breast/ovarian cancer or menopause, and lose their reproductive function iatrogenically.^36-37^ These patients would then not be accounted in studies analyzing serum AMH.

Though there is no uniform normal range for AMH, in general, the mean value of 2.0 ng/ml in gBRCA in our IPD meta-analysis is well below the lower range of age-matched normal (2.9 ng/mL).^38^ Within that range, an average difference of 1.32 ng/ml is highly significant as it is 35% lower than the controls, which could translate into a shortening of reproductive life period by 10 years.^39^

Despite our repeated efforts, we could not obtain raw data from 7 of 12 studies we identified, all involving unaffected woman. Because the data from the non-participating studies greatly varied in data format, availability and/or quality, it was not possible to perform any reasonable meta-analysis or preliminary data processing/standardization as a sensitivity or secondary analysis. However, the 5 studies that were included represent >80% of all published data on affected women. For this reason, our analysis is robust for the relationship between g*BRCA* and ovarian reserve in women who developed breast cancer. However, the non-participation of seven studies that nearly exclusively studied unaffected women does not allow us to reach a firm conclusion on the association of diminished ovarian reserve with g*BRCA* in unaffected carriers.

In conclusion, by individual patient-level data analysis from five centers, we showed that women with g*BRCA* have lower AMH levels compared to those without, and this appeared to be restricted to those with g*BRCA1*. Therefore, based on this IPD analysis and the supporting basic science and translational data,^5,6,40^ we recommend that especially the affected women with g*BRCA1* should proactively receive reproductive and fertility preservation counseling if they are postponing childbearing to the third decade and beyond. This conclusion appears to be firmer for affected women as our IPD meta-analysis predominantly studied those with breast cancer, but further original and meta-analytic studies are needed to determine if there is a difference between the ovarian reserve of affected and unaffected women with *gBRCA* and to understand the magnitude of ovarian reserve differences between women with g*BRCA1* and g*BRCA2*.

The authors declare there is no conflict of interest. Matteo Lambertini acted as a consultant for Roche and Novartis, and received honoraria from Theramex, Takeda, Roche, Lilly, Pfizer and Novartis outside the submitted work.

## Data Availability

The data will be available if the corresponding author permits.

## Contributions

Conception of the idea: KO; Design: KO, VT, HB; Study execution: KO, VT, HB; Provision of study materials: KO, VT, ML, EW, BK, ID, YL, FC; Manuscript writing: KO, VT, HB, ML, ID, EW, BK, YL, FC; Statistical Analysis: HB, VT; Final approval: All authors.

