## Supplementary material for "Association of Germline BRCA Pathogenic Variants with Diminished Ovarian Reserve: A Meta-Analysis of Individual Patient-Level Data": supp material

Fixed Effect (FE) models (commonly used in Econometrics) were chosen as primary method, which may be best suited for a relatively small number of centers and when each center has a clear and specific meaning, rather than ‘5 centers’ being viewed as a random sample of a target population. However, we also used Random Effects (RE) models as secondary/sensitivity analyses based on a reviewer’s recommendation where center was treated as cluster and with ‘random intercept’24,25, and we reached similar results.
