## Supplemental Table 1 for "Association of Germline BRCA Pathogenic Variants with Diminished Ovarian Reserve: A Meta-Analysis of Individual Patient-Level Data"

| Study (Year) | Country | Design | Population | Sample size | Data Reporting Format | Limitations |
| --- | --- | --- | --- | --- | --- | --- |
| Titus S et al. (2013)* | United States | Prospective | Affected with breast cancer | *gBRCA1/2+* (n=24)  *gBRCA*- (n=60) | Mean/SD | Not adjusted for OCP and smoking. |
| Wang ET et al. (2014)* | United States | Cross-sectional | Unaffected | *gBRCA1/2*+ (n=99)  *gBRCA*- (n=54) | Mean/SD | No adjustment for OCP and smoking. |
| Phillips KA et al. (2016) | Australia | Cross-sectional | Unaffected | *gBRCA1*+ (n=172)  *gBRCA1*- (n=216)  *gBRCA2*+ (n=147)  *gBRCA2*- (n=158) | Adjusted % decrease (CI) available in natural log in linear regression in terms of beta coefficient | - |
| Johnson L et al. (2017) | United States | Prospective | Unaffected | *gBRCA1*+ (n=55)  *gBRCA2*+ (n=50)  *gBRCA*- (n=26)  Low risk controls (n=64) | Mean (CI) available for geometric mean, and odds ratio for binary outcome (AMH<1) | Relatively younger study groups. BMI was available for only 41% of the subjects. Affected women and women with RRSO were excluded. |
| Lambertini M et al. (2018)* | Belgium | Retrospective analysis of database from two previous studies | Affected with breast cancer | *gBRCA*+ (n=29)  *gBRCA*- (n=72) | Median and interquartile range | No adjustment for potential confounders+. |
| Michaelson-Cohen R et al. (2014) | Israel | Retrospective | Unaffected | *gBRCA*1/2+ (n=41)  Healthy controls (n=324) | Mean/SD available, along with age-matched mean/SE | No adjustment for potential confounders+. Women with polycystic ovary syndrome were not excluded*, BRCA1 and 2 mutation carriers were analysed collectively. |
| Giordano S et al. (2016) | United States | Prospective | Unaffected | *gBRCA1*+ (n=68)  *gBRCA*- (n=56) | Mean only, no SD | No adjustment for potential confounders+. |
| Van Tilborg TC et al. (2016) | Netherlands | Cross-sectional | Unaffected | *gBRCA1/2* + (n=124)  Non carriers (n=131) | Median/range, relative change (CI) | Relatively young mean age. BRCA1 and 2 mutation carriers were analysed collectively. |
| Ben-Aharon I et al. (2018) | Israel | Prospective | Unaffected | *gBRCA*+ (n=33)  *gBRCA*- (n=15) | % of control, with associated SE. Raw age available (but unpaired with AMH) | No adjustment for potential confounders+. |
| Gunnala V et al. (2019) | United States | Retrospective | Women affected and unaffected with breast cancer | *gBRCA*+ (n=57)  *gBRCA*- (n=738) | Unadjusted mean/SD, and adjusted mean difference (CI) available. | No adjustment for potential confounders+. Assume the controls are in fact BRCA-negative even though they were not tested. Potential selection bias (including patients that underwent fertility preservation and those with severe mutations that underwent RRSO were not taken into account) |
| Son KA et al. (2019)* | South Korea | Retrospective | Affected with breast cancer | *gBRCA*+ (n=52)  Non-carriers (n=264) | Median (interquartile range) or number (percentage) | - |
| Lambertini M et al (2019)* | France | Secondary analysis of prospectively collected samples | Affected with breast cancer | *gBRCA*+ (n=35)  *gBRCA*- (n=113) | Median with interquartile range | No adjustment for potential confounders+. |

Supplemental table 1: Description and limitations of the original studies investigating the association between germline pathogenic *BRCA* variants (g*BRCA*) and serum AMH levels that were targeted for individual patient data collection.

* The studies that provided raw data and were included in the IPD meta-analysis.

PCOS= Associated with spuriously elevated AMH levels

+Potential confounders: Age, body mass index (BMI), OCP use and smoking,

Abbreviations: AMH, Anti-mullerian hormone; OCP, oral contraceptive pill; RRSO, risk reducing salpingo-oophorectomy
